# Many Professions, One Voice: The Role of Language, Recognition and Leadership in Shaping Professional Identity Within the Healthcare Science Professions

**DOI:** 10.1101/2025.09.11.25335393

**Authors:** Cerys Davies, Ffion Wynn, Sarah Bant

## Abstract

**Background:** Professional Identity (PI) significantly influences how Healthcare Science (HCS) professionals perceive their roles and contributions within the NHS. Strong PI is closely linked to improved collaboration, resilience and patient care, yet research on PI among HCS professionals has been limited.

**Objectives:** This study aimed to investigate how HCS professionals in NHS Wales perceive their PI and identify factors that shape and support PI.

**Methods:** A mixed-methods approach was employed. 263 participants answered an initial survey and a further 42 participants attended focus groups to discuss attitudes and experiences in greater depth.

**Results:** Key findings highlight the essential role of professional recognition, the impact of professional bodies, the importance of language and the influence of mentorship and team culture in shaping PI.

**Conclusions:** The findings support recommendations for enhanced involvement of professional bodies, strategic changes in language and terminology, promotion of professional diversity and the development of robust mentorship and local leadership networks.

**KEY MESSAGES:** *What is Already Known on This Topic?:* Healthcare science (HCS) professionals account for approximately 7% of the NHS Wales workforce. These professionals are essential to diagnostic services, patient care, quality management and leadership within the NHS. Previous research has found that professional identity (PI) is closely associated with enhanced job satisfaction, effective interprofessional collaboration, professional resilience and successful career progression. The literature also identifies the positive influence of professional recognition, regulatory structures, mentorship and research activities on PI. However, identity research with HCS professionals is limited, with HCS professionals continuing to experience limited visibility and challenges related to professional status and belonging.

*What this Study Adds:* This study fills a crucial gap in the research on PI among HCS professionals. While previous literature established the general importance of PI for job satisfaction, collaboration, and career progression, identity research specific to HCS professionals has been limited, with many facing challenges around visibility, status and belonging. By addressing the specific experiences and needs of HCS professionals, this research advances understanding of PI beyond broader healthcare contexts and acknowledges the challenges faced by smaller workforce groups who often feel underrepresented and overlooked.

*How this Study Might Affect Research, Practice or Policy:* The findings of this research offer practical directions for enhancing PI among HCS professionals. Strengthening links with professional bodies, revising language to better reflect the diversity and expertise of HCS roles, and building robust mentorship and leadership structures are likely to improve job satisfaction, multidisciplinary work and patient outcomes. Resulting actions have already been incorporated into the workplan for the national HCS workforce transformation programme (the ‘HCS Programme’) in NHS Wales.

## INTRODUCTION & BACKGROUND

Healthcare science (HCS) professionals account for approximately 7% of NHS Wales workforce according to Electronic Staff Record Data (December 2023). This is over 7,000 individuals working across six professional families: Laboratory Sciences and Genomics, Physiological Sciences, Physical Sciences, Clinical Computational Sciences, Radiography, and Operating Department Practice (see Figure 1). These roles are critical to diagnostic pathways, quality management, radiation safety, leadership and patient care, and yet the professions do not necessarily identify or effectively collaborate with each other [1].

**Figure 1:**
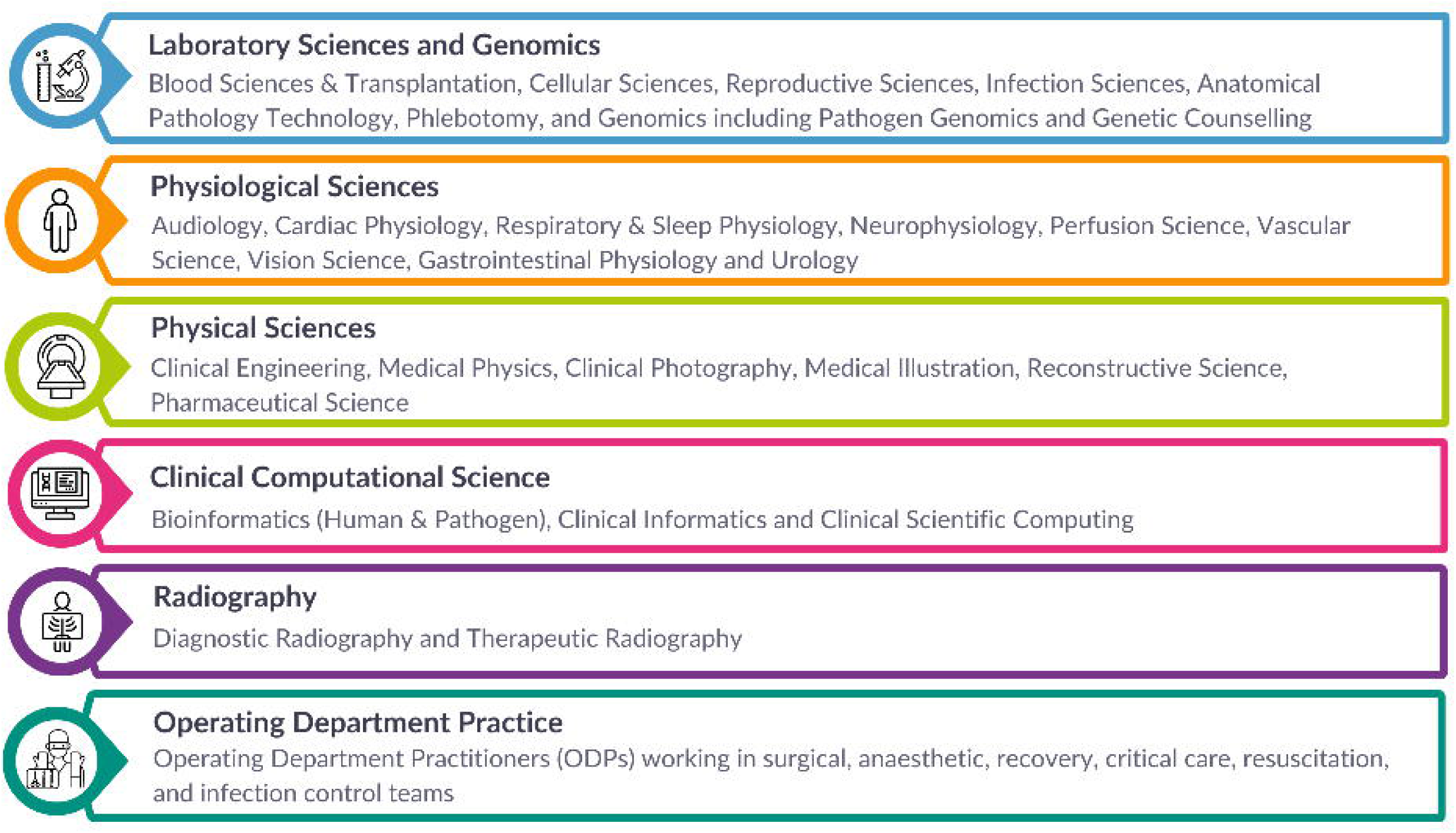
Healthcare Science Professions in NHS Wales

PI is defined as how individuals perceive their role within their profession and the wider healthcare community [2]. Theories such as Social Identity Theory [3], Role Theory [4], Extended Professional Identity Theory [5] and Professional Socialisation Theory [6] explain the development of PI through group membership, role expectations, interprofessional identity and mentorship. Strong PI is linked to job satisfaction, resilience against burnout, career development and effective patient care [7, 8, 9], the fundamentals of each of these theories are illustrated in Figure 2.

**Figure 2:**
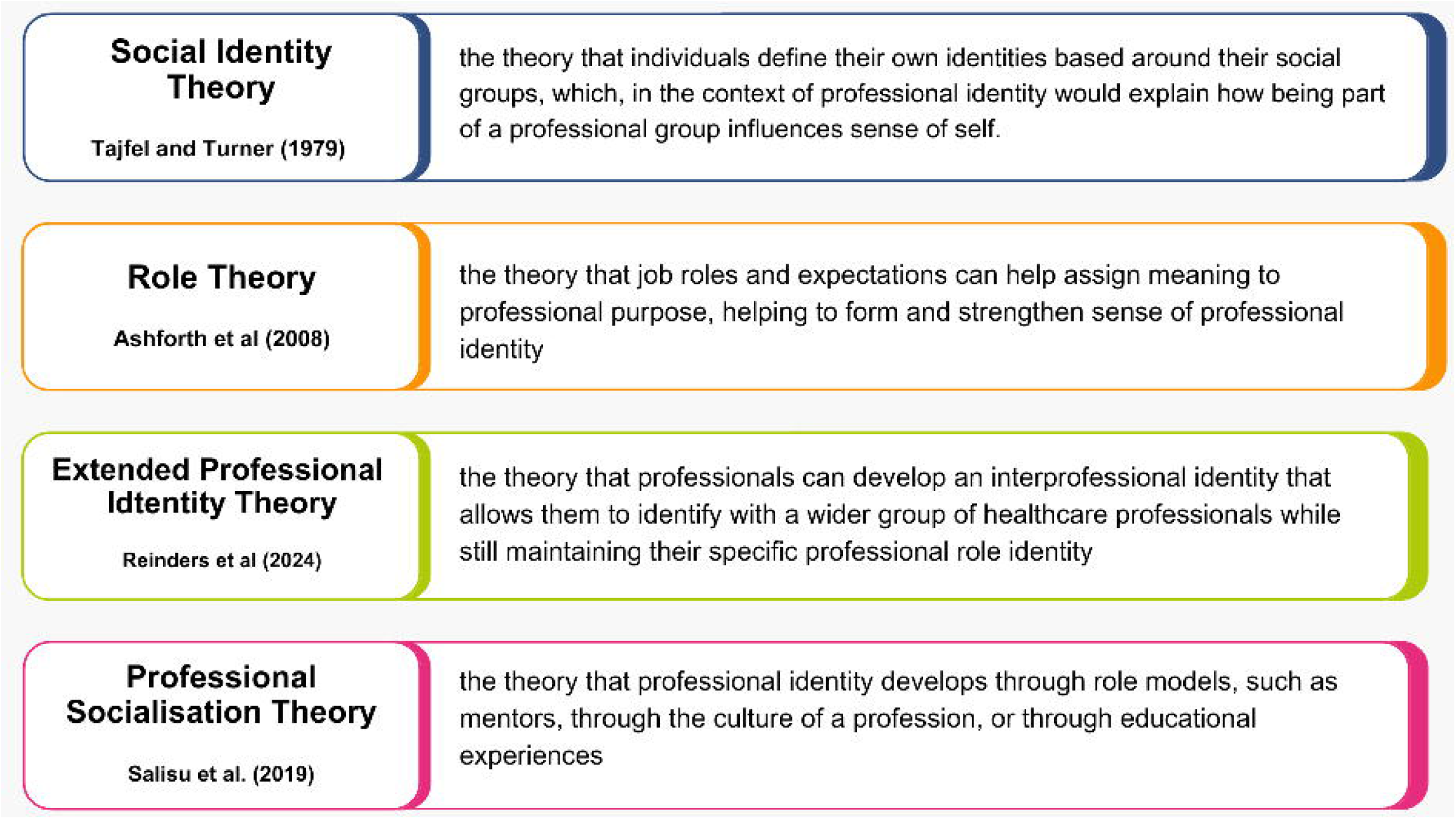
Professional Identity Theories Explained

Research indicates that having a strong sense of PI results in an individual ‘thinking, acting, and feeling’ like their profession [10, 11]. Studies show that mandatory registration enhances professional status [12], while lack of regulation can undermine sense of PI, as seen in Clinical Technologists and Sonographers [13, 14]. Many report that leading in research is important to PI as it improves profession visibility to clinical colleagues, to patients and to the media [15].

The Cavendish Review Ten Years On [16] reports on the perceived invisibility of support staff and assistants in HCS, which can affect sense of belonging and recognition. While some consider themselves as belonging to the HCS community, the focus of belonging is often to smaller communities within their specialties [17].

### AIMS AND OBJECTIVES

This research aims to gather new evidence of perceived PI within the HCS workforce in NHS Wales. Whilst additional issues are likely in Wales, where Diagnostic Radiographers, Therapeutic Radiographers and Operating Department Practitioners (ODPs) also sit under the HCS umbrella, the methodology selected will ensure learning may be drawn for the whole UK.

The umbrella term ‘Healthcare Science’ appears to be widely misused and misunderstood, with this term being used incorrectly to refer to “the science of” professions outside of this umbrella such as nursing, physiotherapy and pharmacy. It is therefore important that people have a clear understanding of what this umbrella terminology aspires to cover, and that we understand how people identify with the terminology and feel included by it.

This study adopts a pragmatic mixed methods approach, valuing both quantitative and qualitative data. The research was conducted in two phases:

1. Quantitative and explorative survey designed to identify patterns and opinions
2. Qualitative phase used to explore the findings in greater depth.

### METHODS: STUDY 1

An initial survey was conducted to explore current perspectives on PI within the HCS workforce in Wales. The objective was to establish the relevance and necessity of further research in this area as existing literature in this area was limited.

#### Sampling and Participants

A total of 263 participants answered the survey. All six professional families were represented: Laboratory Sciences (n=35), Physiological Sciences (n=57), Physical Sciences (n=70), Clinical Computational Sciences (n=1), Radiography (n=60) and ODPs (n=30). A further 10 respondents identified as no longer working in HCS.

#### Data Collection

The literature review helped identify key themes and gaps in existing research, or highlighted nuances specific to the HCS workforce which directly informed the structure and content of the survey.

The survey focused on five key themes:

1. terminology and language used to define PI
2. changing terminology with different audiences
3. registration and protected titles impact on PI
4. connection to the umbrella terminology ‘healthcare science professional’
5. impact of professional misidentification

The survey was shared via the HCS Programme’s social media platforms and newsletters throughout August 2024. Participation was anonymous and voluntary.

#### Data Analysis

Quantitative analysis, described in full below, was conducted on the survey responses to determine a baseline understanding of PI of the HCS workforce in Wales.

### FINDINGS: STUDY 1

#### Connection to Umbrella Terminology

Less than half (46%) of respondents reported feeling included by the term ‘healthcare science professional’. When excluding Diagnostic Radiographers, Therapeutic Radiographers and ODPs, who are included within Allied Health Professionals (AHPs) or Nursing in other areas of the UK, this figure rose to 59%. Responses by professional area are shown in Figure 3.

**Figure 3:**
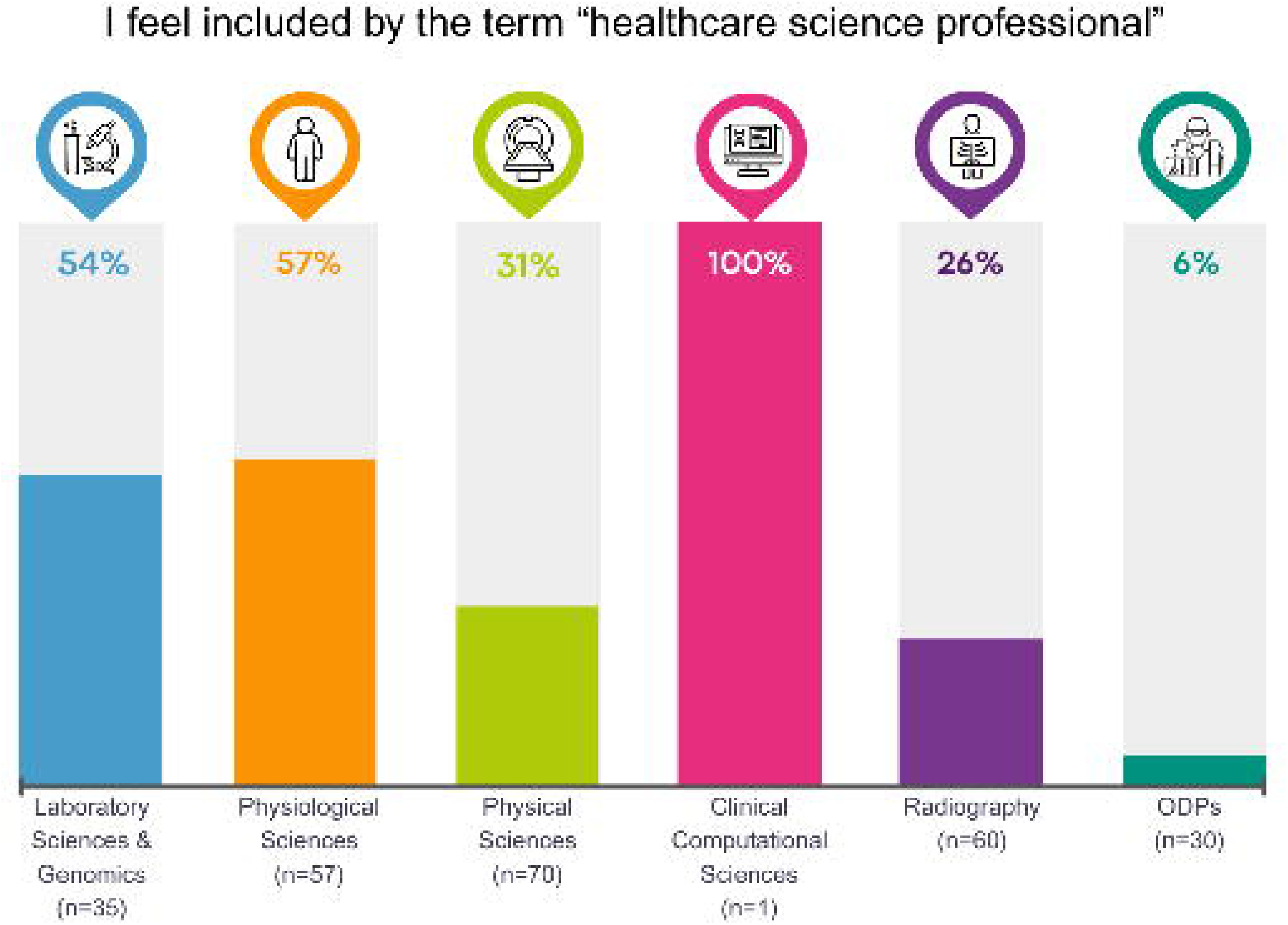
Profession Feelings on ‘Healthcare Science Professional’ as an Umbrella Term

#### Impact of Registration

Registration status and protected titles emerged as a significant factor in PI. Among those on mandatory registers such as with the Health and Care Professions Council (HCPC), 96% considered registration to be essential to their identity. This attitude was also shared by 67% of those on Professional Standards Authority accredited registers, such as with the Academy for Healthcare Science (AHCS).

#### Importance of Recognition

Language was tied to a sense of pride and being valued by colleagues and patients and suggested significant emotive responses when the language used does not align with one’s own PI. This often-left individuals feeling undervalued, frustrated or indifferent (see Figure 4).

**Figure 4:**
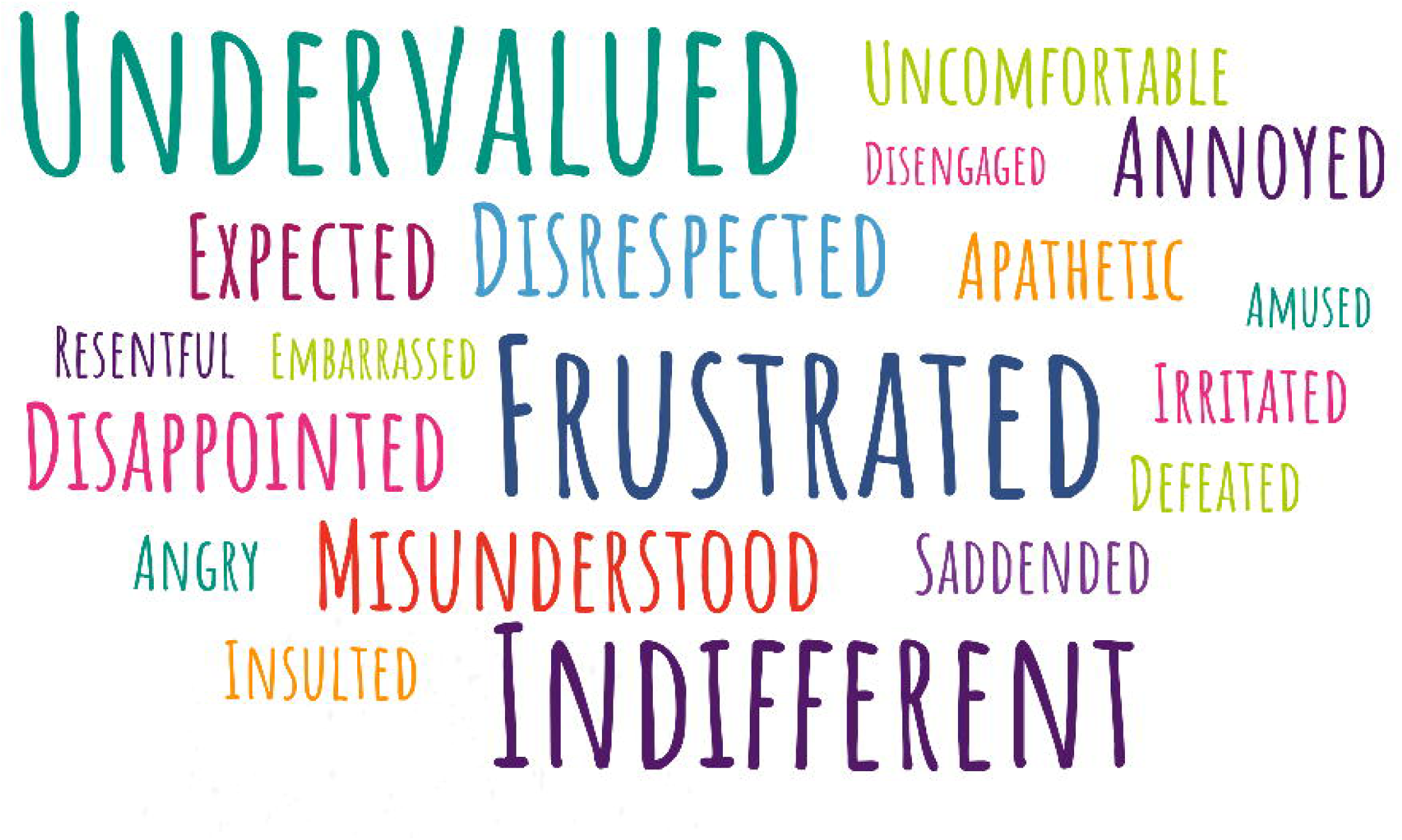
Word Cloud of Emotions when Professionally Misidentified

### CONCLUSION: STUDY 1

The findings highlight a complex and varied understanding of PI within the HCS workforce in Wales. Consequently, a second, qualitative phase of research aimed to explore these views further through profession-specific focus groups.

### METHODS: STUDY 2

Building on the findings from the initial survey, this phase aimed to further contextualise the survey findings, better understand how to increase feelings of inclusion and to identify potential implications for professional recognition.

#### Participants

Participation in the focus groups was voluntary and open to all HCS professionals. A total of 42 individuals participated: Laboratory Sciences (n=6), Radiography (n=9), Physical Sciences (n=8), Physiological Sciences (n=9), and ODPs (n=10).

#### Data Collection

A series of profession specific focus groups were held via MS Teams. Ripple Effects Mapping (REM) was used to facilitate the focus groups [18]. REM was chosen for the focus groups to encourage open, participant-led reflection and conversation, enabling deep insights into PI through collaborative visual mapping of experiences.

The researcher started the focus groups by welcoming the attendees and setting ground rules for open and honest communication, the purpose of REM, and how the focus group would run.

Participants first asked each other set questions in break out rooms away from the researcher and facilitator. Allowing participants to discuss freely in breakout rooms before group mapping is crucial in REM, and encourages honest and authentic discussions:

- How do you describe your PI?
- Describe a time you felt proud of your PI.
- Where do you feel your sense of PI came from?
- Has your sense of PI ever changed/evolved?

Participants were then brought back to the main room to report back on their discussions, and the feedback was recorded. The facilitator ensured each member had chance to express their views and experiences, as the researcher visually mapped out the conversation via MS Whiteboard.

#### Data Analysis

Reflexive Thematic Analysis (RTA) [19] was used to draw out initial themes from transcripts. Focus groups were recorded and transcribed, the recording re-watched to gather deeper understanding and check that all transcripts were accurate. The coding procedure involved identifying recurring themes in the transcripts and selecting impactful participant quotes for illustration. Researcher bias was mitigated by using REM, enabling participants to collectively agree that the findings accurately reflected group perspectives.

#### Ethical Approval

Study 2 has been approved by the Cardiff University School of Medicine Research Ethics Committee (REF 24/72).

### FINDINGS: STUDY 2

RTA identified three overarching themes, each with two sub-themes (see Figure 5). These reflect shared and profession-specific experiences, shaped by recognition, language, regulation and professional development.

**Figure 5:**
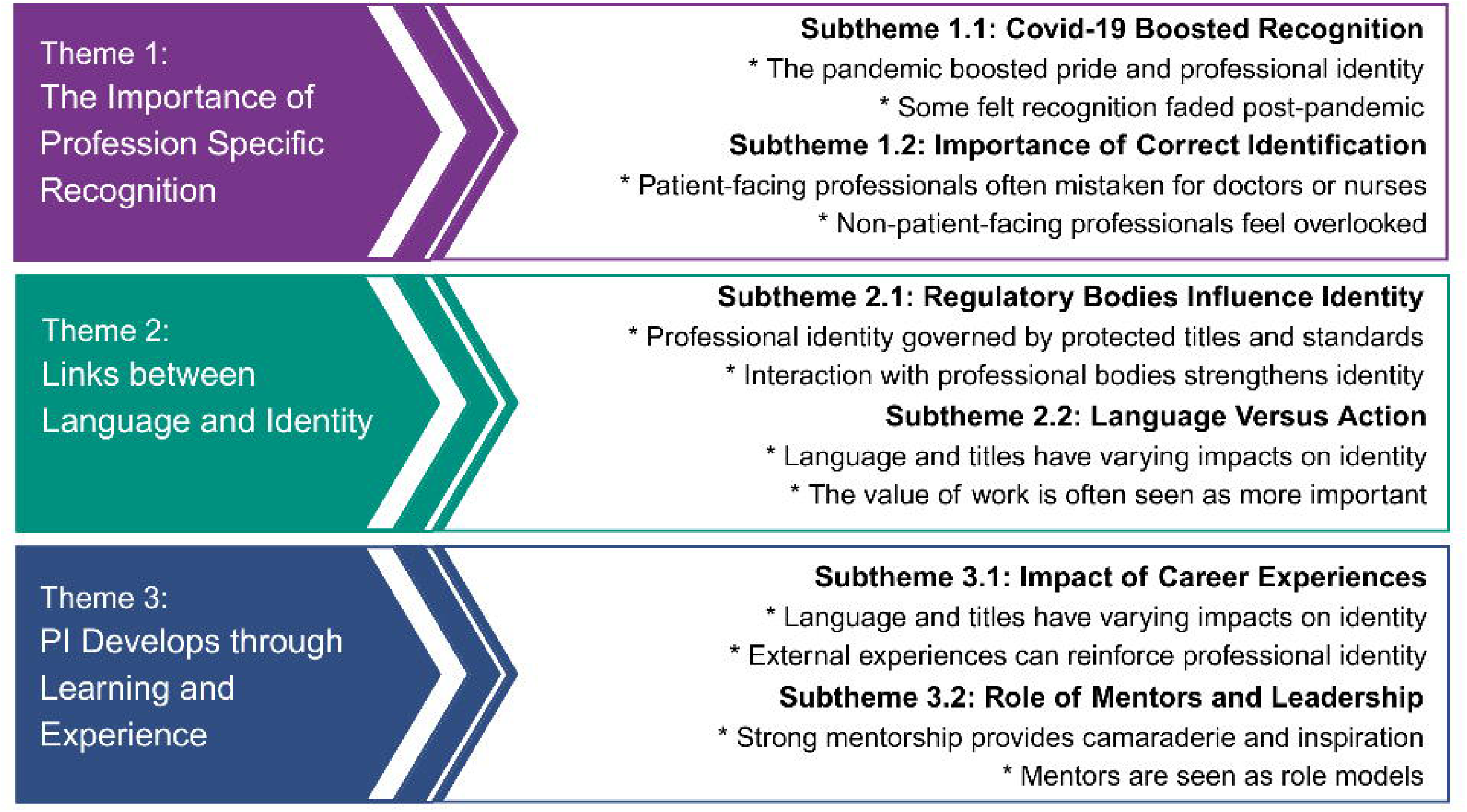
Summary Of Themes and Sub-Themes from Focus Groups.

### Theme 1: The importance of profession specific recognition in strengthening sense of identity

#### Subtheme 1.1: Covid-19 boosted recognition of the profession

The pandemic significantly impacted the NHS, influencing many professionals’ identities. ODP focus group members stated it felt like a “massive journey” (ODP07) that brought new opportunities and lasting positive effects to their field.

> “Covid was a turning point for us as a profession within our Health Board as well because, off the back of that, we then had our first ODPs in ICU working alongside the nurses within the same capacity”. (ODP06)

For many, the pandemic was the first time that they felt “seen” (DR01) in their profession, leaving them with a “warm and fuzzy feeling” (BMS02). Healthcare professionals and the public acknowledged the vital role of the HCS workforce:

> “It took something like Covid where people realise that we actually are entirely essential to the service and that it doesn’t really function without us”. (DR06)

Some professions, particularly within the physiological sciences, felt that this recognition for the profession faded as the pandemic became less prevalent:

> “[…] in respiratory, Covid was probably a time I felt proud of our profession because suddenly people started to realise that we exist and what we could do […] I think people think that with the pandemic leaving, so did respiratory physiologists, because now we don’t exist again”. (CP05)

#### Subtheme 1.2: It is important for others to get my profession correct

Many patient-facing professionals reported being mistaken. One participant shared “a lot of patients just assuming that we’re, you know, nurses or doctors or something” (ODP04).

Another noted “a number of times my colleagues have been called nurses” (CP06), which highlights that misidentification is persistent.

This likely stems from a general lack of public awareness about the range of specialised roles within the NHS, causing many HCS professionals to feel overlooked. For non-patient-facing areas of HCS, this lack of understanding of roles beyond doctors and nurses left them feeling completely invisible:

> “Patients think that the doctors run a test and get the results, and they’re just not aware of like, a whole group of people”. (BMS04)

Some noted that, with regular correction, patients began using the correct titles and even corrected others. This recognition for their work brought a strong sense of pride and validation:

> “When you actually get recognised as a Therapy Radiographer, you know, you kind of beam a little bit because you realise people are taking it on board”. (TR01)

The importance of recognition in shaping and affirming identity is paramount:

> “Recognition is key to identity, and I think we are more recognised than we were, but there’s still a long, long way to go”. (MPCE05)

### Theme 2: Strength of language linked to identity

#### Subtheme 2.1: Regulatory bodies influence identity

PI was strongly governed by the protected titles and standards of professional bodies. Protected HCPC registration titles, such as Biomedical Scientist or Radiographer, define how professionals identify and uphold standards daily. This sense of identity is reinforced through their interactions with professional bodies.

> “Our identity was born out of the way that we were trained and the way that we grew as radiographers, particularly the way that we interact with our professional body and how they identify us”. (DR03)

When discussing language associated with PI, changes in terminology were easily accepted when coming from the regulatory body:

> “When it was CPSM [Council for Professions Supplementary to Medicine], which was the old regulatory body, you were an MLSO [Medical Laboratory Scientific Officer]. I easily transitioned over to biomedical scientist because that is what the new regulatory body said you are. The regulatory body is never going to say you are healthcare scientists, so there’s no identity to that particularly”. (BMS01)

Clinical Scientists appeared to be the protected title with the most flexibility, likely due to the nature of this protected title stretching across a multitude of HCS disciplines. Those who were registered as a Clinical Scientist found that when they identified with this language it sometimes led to misunderstanding or a misconception of their role by others. Protected HCPC titles, such as Biomedical Scientist or Radiographer, define how professionals identify and uphold standards daily.

> “I try and use “clinical scientist” or “healthcare scientist” more on a day-to-day basis. But what I do tend to find then is that people kind of think that you work in a lab because you use the word scientist”. (CP04)

Diagnostic and Therapeutic Radiographers felt particularly detached from the term “scientist”, stating “the term ‘healthcare sciences’ sounds quite cold, it doesn’t sound patient centred” (DR04).

#### Subtheme 2.2: Language versus Action

Many focus group attendees felt language and titles had little to no impact on their sense of PI. The most important thing to some was the value in the work that they do, with one participant stating, “It doesn’t really matter what we are called, it’s what we do that’s important” (CP06).

Even when groups have a strong PI tied to a protected title, their core values reinforce their commitment to maintaining professional standards.

> “Professional identity comes from what you do in the lab” (BMS03)

While titles may not shape identity for all, they do carry legal and functional implications in some healthcare science roles. For instance, “radiographers are able to give certain drugs because they’re on the HCPC [compared to clinical technologists]” (MPCE06).

For some, PI came from the joy and pride they felt doing their job, and this wasn’t affected by a lack of understanding from others:

> “My professional identity doesn’t come from how other people perceive me, you know? I know what I do. I enjoy what I do”. (DR06)

Others believed PI developed from mutual understanding with patients. Patients’ perceptions of professional roles can shape PI, when patients understand these roles clearly it reinforces sense of pride and value.

> “What do they [the patient] think we should be called, I wonder? It’s their understanding of us which is important, isn’t it?” (CP08)

### Theme 3: Professional identity develops through learning and experience

#### Subtheme 3.1: The impact of career experiences

Many noted how personal experiences helped shape PI:

> “When you start independent working, I think that does hit you. The patient is at the end of it, you’re now responsible for everything you do and everything you do not do, every choice you make is on you”. (BMS01)

Being “part of a community” (DR04) provides clarification on how you as an individual fit into the wider pathway, providing a role and contextualising your identity:

> “I think for me, my professional identity didn’t come until a couple of years into the profession where I actually started working in a much bigger oncology department than what I’d started seeing the importance of the role”. (TR01)

External experiences can impact sense of self. Some participants described using their expertise outside the hospital, such as two ODPs who assisted at accident scenes thanks to their specialised skills. Both expressed pride in being taken seriously for their professional background and valued making a difference.

> “I was at a sporting event and one of the players had an injury […] The first aider didn’t have a clue […] When the paramedics arrived, I told him I was an ODP, and that’s when I truly realised how appreciated we were because the paramedics were so glad that I knew what I was doing”. (ODP02)

> “I was at the scene where a car reversed over a gentleman, and he was on the floor in pain. The other witnesses were trying to get him up, I had to tell them ‘no, we’ve got to keep him still’”. (ODP01)

#### Subtheme 3.2: The role of mentors and leadership

Key mentors help shape PI through values, context and experience. Participants recalled that mentors often captured the profession’s essence.

> “I think part of my sense of professional identity was seeing that mentor and how they acted and interacted with patients, they were definitely people that I thought that’s how I want to be”. (CP05)

Strong mentorship and support from the wider team provided “camaraderie” (ODP08) and was seen as essential to PI. Mentors and other senior staff were seen as a source of “inspiration” (BMS01) to many:

> “I think for me personally, a lot of it came from the manager in the lab when I first started. He really walked the walk, he talked the talk […] he had a great way of putting it all in context, which probably instilled that origin of the pride in me more than anything else”. (BMS02)

### DISCUSSION: STUDY 2

The pandemic highlighted strengthened the PI of HCS staff in the NHS. Many felt recognised and valued, particularly ODPs whose roles expanded during the crisis, though this visibility wasn’t constant across all specialisms.

Regulation also shaped the sense of PI, with protected titles offering clarity and supporting ethical practice. Comparisons between disciplines, such as Clinical Technologists and Radiographers, reflected ongoing concerns about recognition and scopes of practice.

Mentorship played a strong role in fostering pride, purpose, and leadership skills, helping professionals feel connected and empowered. Overall, recognition, regulation, and mentorship were key factors in building confidence and leadership within HCS in Wales.

#### Study Strengths and Limitations

This study provides novel insights into the PI of the HCS workforce in Wales, an area where existing research is limited and the need for collaboration, the coming together with “one voice” is key to realising the potential for these essential professions. It includes a wide range of perspectives from both patient-facing and non-patient-facing roles, as well as professions with varying degrees of public recognition.

A notable strength is its two-phase mixed-methods design, with the initial survey effectively informing subsequent focus group discussions.

Whilst the sample size was appropriate for the qualitative approach (n=42), certain disciplines were underrepresented, which restricts the generalisability of the findings. Focus group dynamics may have introduced bias due to dominant voices or social desirability; however, the use of REM ensured discussions were open and reflective, mitigating potential power imbalances. Nevertheless, individual interviews may have yielded more in-depth insights.

RTA enhanced the depth of analysis but also introduced potential for researcher subjectivity. This risk was balanced through the process of REM, which allowed for real-time confirmation of themes.

The research also highlights how language influences PI. Some participants noted a sense of detachment from umbrella terms like ‘Healthcare Science Professionals’, however this is not a term that can be changed.

## CONCLUSION

This study offers new insights into the PI of the HCS workforce in Wales, highlighting how recognition, regulation and mentorship shape how professionals see themselves and their roles within the NHS. The findings suggest that, whilst umbrella terminology may have its use, the individual identity of professions within should be valued.

### Key Findings

- Recognition during the pandemic affirmed PI for many, though this was not sustained across all specialisms
- Regulatory bodies and protected titles significantly influenced PI
- Mentorship played a critical role in building confidence, pride and leadership aspirations
- Language and terminology were important, though some professionals felt disconnected from umbrella terms
- Public and peer misidentification remains a barrier to PI and visibility.

#### Implications for Leadership

These findings have significant implications for leadership across NHS Wales and the UK. Strong PI underpins confidence, clarity and collaboration, all key to effective leadership.

Leaders are essential in supporting identity development, including:

- Using inclusive, accurate language that reflects the protected titles and professional affiliations of HCS workforce
- Promoting visible role models from across the HCS professions to inspire early-career professionals and reinforce a sense of belonging
- Embedding mentorship and leadership development into workforce strategies, recognising the formative role of mentors in shaping PI
- Ensuring strategic visibility of HCS roles in organisational communications, planning and recognition initiatives.

Recent research with AHP identity supports these recommendations. Eddison et al. [20] point out the limited representation of some AHP groups in senior NHS leadership roles, noting that leadership positions are often dominated by a narrow range of professions. Similarly, Mizzi and Marshall [21] highlight barriers to leadership development for early- and mid-career AHPs, underscoring the need for equitable access to leadership pathways.

Given that HCS professionals occupy only 30% of the 21 Directors, Deputies and Assistant Directors of AHP and Health Science roles across NHS Wales health boards, despite the roles covering both HCS and AHP, it is crucial to promote and support HCS professionals to ensure balanced representation and leverage their expertise for improved healthcare outcomes. By creating a culture that values and affirms PI, leaders can enhance workforce engagement, retention, and leadership capacity, ultimately supporting the delivery of safe, person-centred care.

#### NEXT STEPS

To strengthen PI, the study recommends actions such as public education, improved communication, mentorship, clear career pathways, consistent terminology and advocacy for protected titles. These measures aim to enhance a unified and visible identity for HCS professionals within the NHS.

## Data Availability

All transcripts produced in the present study are available upon reasonable request to the authors

## Notes

### Competing Interest Statement

The authors have declared no competing interest.

### Funding Statement

This study did not receive any funding.

### Author Declarations

The School of Medicine Research Ethics Committee or Cardiff University gave ethical approval for this work.

